# Differences in Neuropsychiatric Features in Black and White Individuals Diagnosed with Frontotemporal Degeneration

**DOI:** 10.1101/2023.01.27.23284692

**Authors:** Hannah Jin, Corey T. McMillan, Isabel Yannatos, Lauren Fisher, Emma Rhodes, Sarah F. Jacoby, David J. Irwin, Lauren Massimo

## Abstract

**Background:** Neuropsychiatric symptoms are highly prevalent in frontotemporal degeneration (FTD). Prior research suggests there are disparities in the clinical presentation of dementia when comparing Black and White individuals but this has not been investigated in the context of FTD specifically. The aim of the present study is to investigate racial disparities in dementia severity, functional impairment and neuropsychiatric symptoms in individuals with a clinical diagnosis of FTD.

**Methods:** Using National Alzheimer’s Coordinating Center (NACC) data, we evaluated 63 Black and 2,356 White individuals living with a clinical diagnosis of FTD (behavioral variant FTD or primary progressive aphasia) and a healthy control group of 1899 Black and 9122 White individuals with normal behavior and cognition. We compared demographic characteristics, dementia severity, functional impairment and neuropsychiatric symptoms at initial NACC visit by examining differences on Clinical Dementia Rating Scale (CDR), Functional Assessment Scale (FAS) and Neuropsychiatric Inventory (NPI) using multivariable linear and logistic regression models, covarying for age at visit, disease duration, sex, and education. Models evaluating differences in neuropsychiatric symptoms additionally controlled for dementia severity.

**Results:** Black individuals with FTD were considerably underrepresented, comprising only 2.5% of the clincial sample. In comparison to White individuals, Black individuals with FTD had a higher degree of dementia severity on CDR (CDR sum of boxes; p=0.03 CDR global, p=.006), greater functional impairment (FAS total; p=0.01) and more delusions (p=0.01), agitation (p=0.04) and depression (p=0.03) on the NPI. White individuals with FTD were more likely to demonstrate apathy (p=0.03). Within the normal control group, Black individuals reported less depression, anxiety, agitation, and nighttime behaviors (depression; p < 0.001; anxiety; p = 0.006; agitation; p = 0.048; nighttime behaviors; p < 0.001) compared to White normal controls.

**Discussion:** The present study suggests there are significant disparities in dementia severity, functional impairment and neuropsychiatric presentations at initial visit between Black and White individuals with FTD. Future work must address racial disparities in FTD and their underlying social determinants as well as the lack of representation of non-White individuals in nationally representative neurodegenerative disease registries in order identify appropriate interventions.

## INTRODUCTION

Frontotemporal degeneration (FTD) is a progressive neurodegenerative disease that affects social comportment, executive function, as well as speech and language.^1^ Neuropsychiatric symptoms are exceedingly common in FTD and are a major source of distress for caregivers of persons with FTD^2,3^ In the United States, the rate of dementia is highest in racially minoritized populations;^4^ however, there is a dearth of literature on racial differences in FTD, a common form of young-onset dementia. Prior studies have compared neuropsychiatric symptoms in Black and White individuals with dementia, but patient samples have typically either been comprised of patients with Alzheimer’s Disease (AD)^5,6^ or were heterogenous dementia samples that were not stratified by dementia type.^7,8^ This body of work has demonstrated that compared to White individuals, Black individuals are more likely to show psychotic symptoms,^5,8,9^ particularly hallucinations,^6,7,10^and present with a greater degree of functional impairment at the time of an initial visit,^11^ highlighting differences in clinical presentation by race in persons with dementia. However, it is unclear if the differences in neuropsychiatric symptoms that emerge when comparing racial groups are specific to neurodegenerative disease or reflect differences and expressions of psychiatric symptoms in the general population;^12^ as prior research investigating racial disparities in the symptomotology associated with dementia have not included a control group of adults with normal cognition. With recognition that racial differences in FTD symptomatology are not biologically driven, the socioeconomic, structural and cultural factors associated with racial discrimination have the potential to impact clinical presentations and diagnostic recognition of FTD. Therefore, the overall purpose of this study was to examine differences in neuropsychiatric symptoms and functional presentations comparing Black and White with a diagnosis of FTD. Specifically, we used National Alzheimer’s Coordinating Center (NACC) data to: 1) determine whether neuropsychiatric symptoms differ when comparing Black and White individuals clinically diagnosed with FTD; 2) determine whether neuropsychiatric symptoms differ when comparing Black and White individuals with normal cognition; and 3) determine if there are differences in functional ability between Black and White individuals clinically diagnosed with FTD.

## METHODS

### Study design and participant selection

We performed a retrospective study of NACC data obtained from self-reported Black and White individuals with a clinical diagnosis of behavioral variant FTD (bvFTD) or a primary progressive aphasia (PPA). NACC is public dataset established in 1999 by the National Institute on Aging that collects standardized clinical and neuropathological data^13^ from Alzheimer’s Disease Research Centers (ADRCs) across the United States, and the data used in this study were collected from June 2005 to August 2021. Due to limited cases, we excluded all other reported races, including American Indian or Alaska Native (N = 4), Native Hawaiian or Other Pacific Islander (N = 3), Asian (N = 62), Other (N = 13), Unknown (N = 24). We define initial visit as the first visit for which each patient had a clinical diagnosis of FTD (i.e., either bvFTD or PPA).^i^ We selected cases who had the following complete outcome data: CDR® Dementia Staging Instrument plus NACC FTLD Behavior & Language Domains, Neuropsychiatric Inventory–Questionnaire (NPI-Q), and demographic variables including race, age, sex, education, and onset of cognitive decline. Disease duration was calculated as time from the onset of cognitive decline (e.g., NACC variable DECAGE) until the initial visit date. We excluded any participant whose disease duration at initial visit was greater than four standard deviations above the mean disease duration (> 239.9 months). Our final sample was comprised of a total of 2,356 White individuals and 63 Black individuals (see study flow diagram in Figure 1). In our final sample, no Black individuals and 2.8% of White individuals reported Hispanic ethnicity. Table 1 summarizes demographics of the FTD group.

**Table 1:** Demographic and clinical characteristics of NACC participants with bvFTD or PPA

|  | bvFTD |  | PPA |  | bvFTD or PPA |  | p<sup>†</sup> |
| --- | --- | --- | --- | --- | --- | --- | --- |
|  | White<br>(N = 1210) | Black<br>(N = 37) | White<br>(N = 1146) | Black<br>(N = 26) | White<br>(N = 2356) | Black<br>(N = 63) |  |
| SEX = F (N<br>(%)) | 456 (37.7) | 20 (54.1) | 568 (49.6) | 21 (80.8) | 1024<br>(43.5) | 41 (65.1) | 0.001 |
| AGE (mean<br>(SD)) | 63.96<br>(10.01) | 65.19<br>(11.04) | 66.58 (8.64) | 68.62<br>(7.79) | 65.24<br>(9.46) | 66.60<br>(9.91) | 0.28 |
| EDUCATION<br>in years (mean<br>(SD)) | 15.28<br>(3.14) | 13.57<br>(4.21) | 15.78 (2.78) | 14.15<br>(3.06) | 15.52<br>(2.98) | 13.81<br>(3.76) | <0.001 |
| DISEASE<br>DURATION<br>in months<br>(mean (SD)) | 70.53<br>(39.62) | 62.94<br>(34.45) | 64.74<br>(30.89) | 66.96<br>(29.54) | 67.72<br>(35.75) | 64.60<br>(32.32) | 0.45 |
| CDRGLOB<br>(mean (SD)) | 1.29 (0.79) | 1.57<br>(0.84) | 0.83 (0.72) | 1.02 (0.91) | 1.07 (0.79) | 1.34 (0.91) | 0.007 |

† To compare demographic data between Black individuals and White individuals, t-tests assuming unequal variance (for continuous variables) or chi-square tests (for categorical variables) were performed

**Figure 1.**
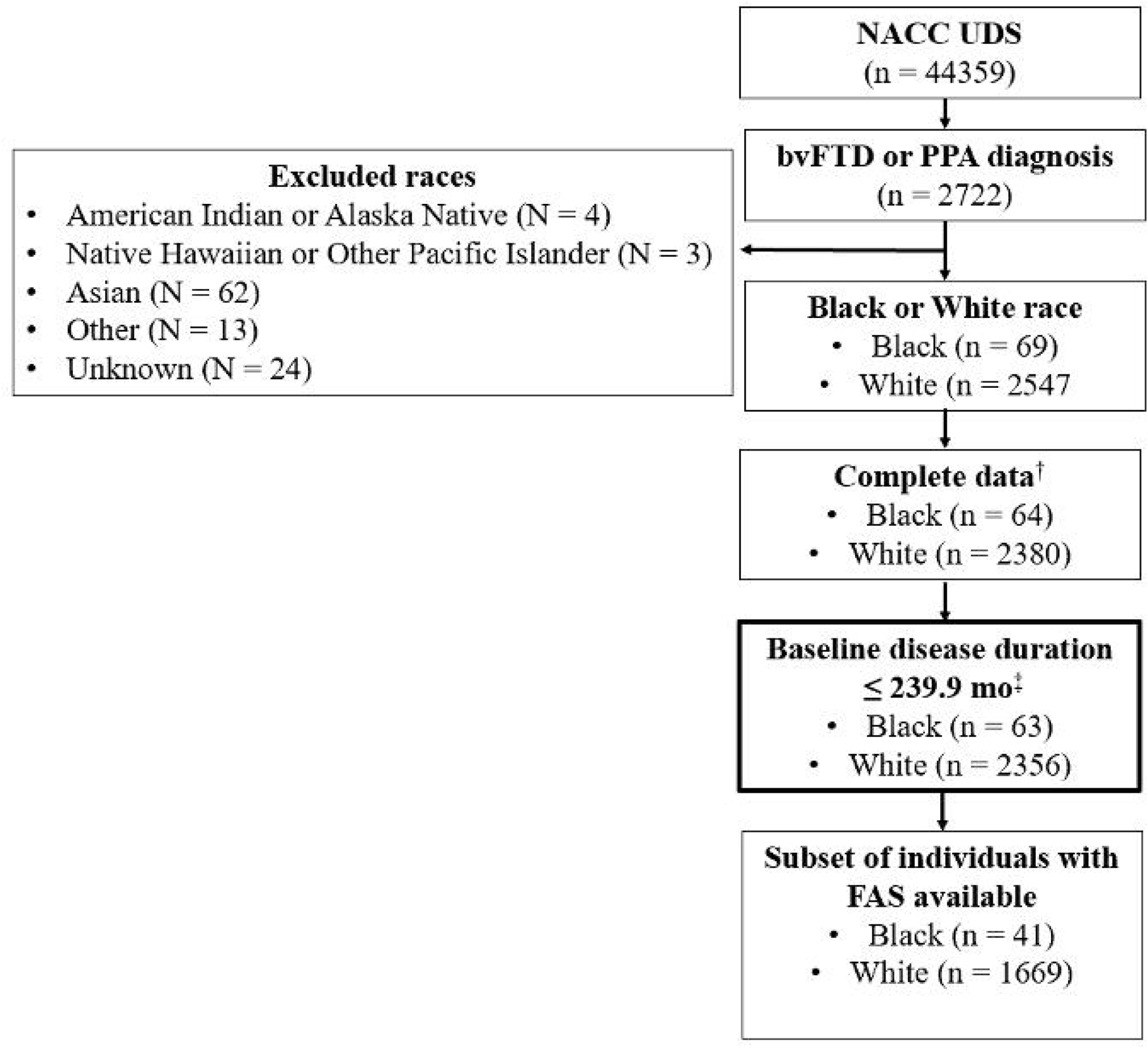
Flow diagram for FTD group. Notes: † “Complete data” refers to non-missing data in the following NACC variables: DECAGE, DEL, HAT X, AGIT, DEPD. ANX, ELAT, A PA. DISN. TRR, MOT. NITE, APP. DELSEV, HALLSEV, AG1TSEV, DEPDSEV, ANXSEV, ELATSEV. APASEV, DISNSEV, IRRSEV, MOTSEV, NITESEV, APPSEV, CDRSUM. CDRGLOB. RACE, AGE. SEX, EDUC, VISITMO. VISITDAY, VISITYR. ‡ 239.9 mo is 4 standard deviations above the mean baseline disease duration (69.9 mo).

To evaluate specificity of neuropsychiatric symptoms in FTD, we conducted an analysis of NPI among a normal control (NC) group. Participants met the criteria for the NC group if they had normal cognition and normal behavior at initial visit (e.g., NACC variable NORMCOG = 0) and maintained a CDR global score of 0 throughout all NACC visits (see flow diagram in Figure 2). Therefore, the control group consisted of 1899 Black and 9122 White individuals. Table 2 describes control group demographics. Qualified researchers may obtain access to all de-identified data in the NACC registry used for this study (http://www.naccdata.org).

**Table 2:** Demographic characteristics of healthy control participants

|  | White<br>(N = 9122) | Black<br>(N = 1899) | p |
| --- | --- | --- | --- |
| SEX = F (N (%)) | 5899 (64.7) | 1475 (77.7) | <0.001 |
| AGE (mean (SD)) | 68.71 (11.20) | 69.22 (8.34) | 0.02 |
| EDUCATION<br>in years<br>(mean (SD)) | 16.15 (2.72) | 14.89 (2.95) | <0.001 |

**Figure 2.**
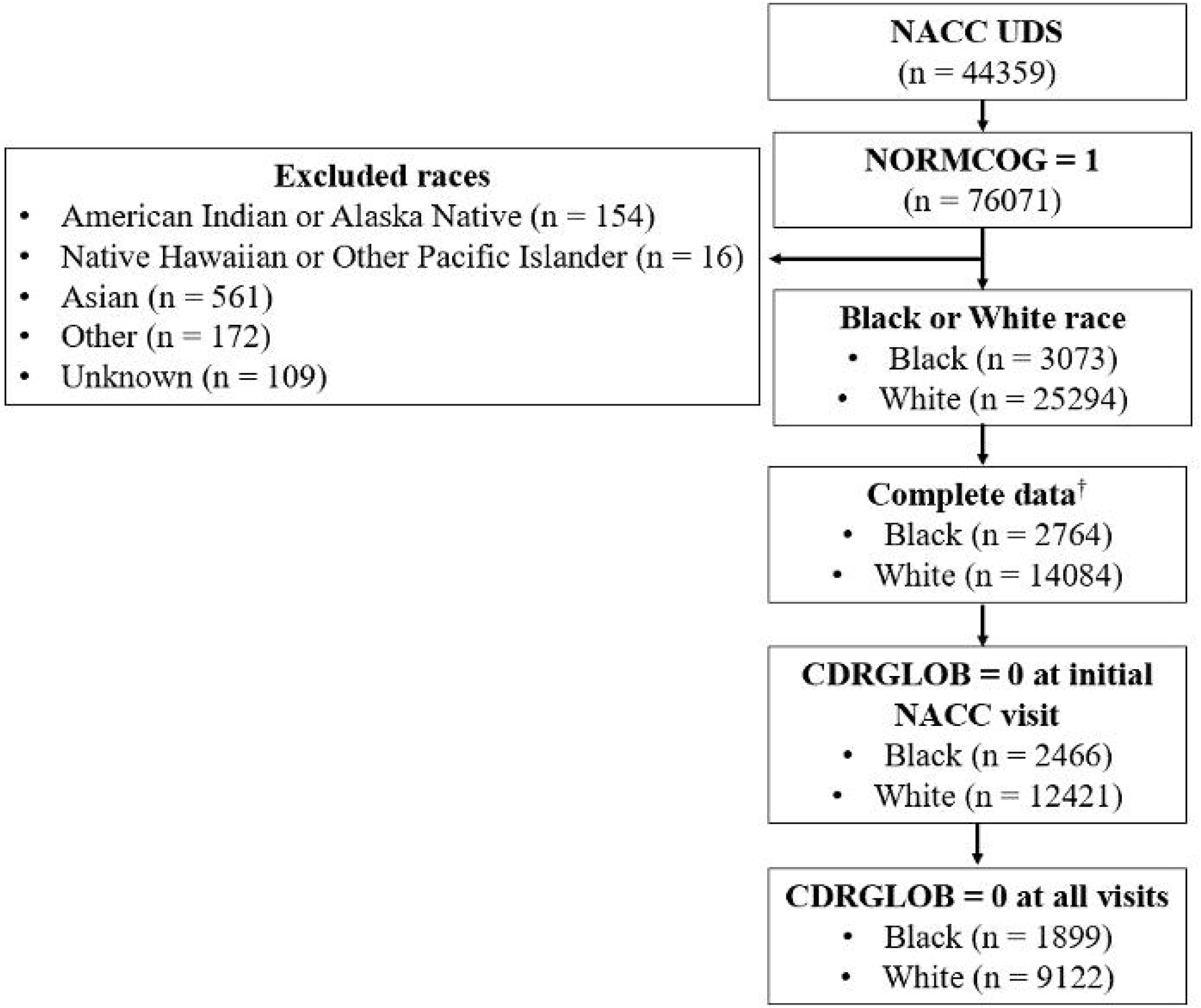
Flow diagram for selection of normal control group. † “Complete data” refers to non-missing data in the following NACC variables: DEL. HALL, AGIT, DEPD. ANX, ELAT, APA. DISN, IRR, MOT, NITE, APP, CDRGLOB, RACE. AGE. SEX. EDUC

### Standard Protocol Approvals, Registrations, and Patient Consents

All contributing ADRCs are required to obtain informed consent from their participants or caregivers and study procedures are approved by local Instituitional Review Boards prior to submitting data to NACC.

### Functional and Behavioral Outcomes

The CDR® Dementia Staging Instrument plus NACC FTLD Behavior & Language Domains evaluates cognitive/functional ability in dementia patients. It consists of eight domains: memory, orientation, judgment and problem solving, community affairs, home and hobbies, personal care, language, and behavior. Based on semi-structured interview with the patient and a knowledgeable informant, category domains are rated as 0 (normal), 0.5 (questionable impairment), 1 (mild impairment), 2 (moderate impairment), 3 (severe impairment). The CDR sum of boxes (CDR SOB) score is the sum of ratings across domains. The global CDR (i.e., CDR global) ranges from 0 (normal), 0.5 (questionable impairment), 1 (mild impairment), 2 (moderate impairment), 3 (severe impairment), indicating the stage of dementia.

The NACC Functional Assessment Scale (FAS) evaluates the patient’s functional ability. This 10-item scale evaluates patient ability to perform daily activities such as preparing a balanced meal. Functional abilities are rated as 0 (normal), 1 (has difficulty, but does by self), 2 (requires assistance), 3 (dependent), not applicable/never did, or unknown. For these item-wise analyses, “Normal,” “Has difficulty, but does by self,” and “Requires assistance” were coded as “0” and “Dependent” was coded as “1” for each individual FAS item. Ratings across the ten items can be summed to yield a FAS total score ranging from 0 to 30.^14,15^

The Neuropsychiatric Inventory (NPI) assesses psychopathology in dementia through a structured interview with the participant’s care partner. NACC collects and reports the Neuropsychiatric Inventory–Questionnaire (NPI-Q), a brief version of the NPI, which can be completed in clinical practice settings and has been cross-validated with the standard NPI.^16^ The NPI-Q assesses twelve behavioral domains: delusions, hallucinations, agitation, depression, anxiety, apathy, irritability, elation, disinhibition, aberrant motor behavior, night-time behavior disturbances, and appetite abnormalities. For each of these twelve neuropsychiatric symptoms, the presence or absence of symptoms in the past month are recorded. If neuropsychiatric symptoms are present, the care partner rates the severity of the behavior as 1: mild, 2: moderate, or 3: severe (e.g., NPI-Q behavioral domain severity score). Severity scores across behavior domains can be summed to yield an NPI-Q total severity score ranging from 0 to 36. Similarly, the number of total symptoms (e.g., NPI Total) present can be calculated from summing the number of domains for which the caregiver responds “yes” to affirm the presence of a neuropsychiatric symptom.

### Statistical Analysis

Analyses were performed with R Studio version 4.1.0. Continuous variables are summarized with mean and SD, and two-sample t-tests were performed to assess group differences in demographics. Chi-square tests of independence were used to examine the distribution of sex.

Linear regression was used to examine the relationship between race and CDR SOB and ordinal logistic regression was used to examine the relationship between race and CDR global. We used linear regression to examine the relationship between race and FAS total, and binary logistic regression to examine the relationship between race and each FAS item among participants with available data (Black n = 41, White n = 1669, see study flow diagram Figure 1 for detailed sample sizes). All CDR and FAS models were adjusted for age, sex, education, and disease duration in months.

Logistic regression evaluated the relationship between race and the presence of each of the 12 neuropsychiatric symptoms and ordinal logistic regression was used to examine the relationship between race and NPI-Q symptom severity of each of the 12 neuropsychiatric symptoms. Linear regression evaluated the relationship between race and total NPI-Q and total NPI-Q severity. In NPI analyses in the FTD sample, we adjusted for age, sex, education, and CDR global as a measure of disease severity. We adjusted for the same covariates in the normal control sample with the exception CDR since all normal controls had a CDR global score of 0.

## RESULTS

The mean age at initial visit in total clinical sample (n=2,419) of Black and White individuals with FTD was 65.3 (SD=9.5) years old and the mean disease duration at initial visit was 67.6 (SD= 35.7) months. As shown in Table 1, age at initial visit and disease duration did not differ between Black and White individuals with FTD. However, Black individuals with FTD were more likely to be female (*x*^2^=10.8, df = 1, p=.001) and had fewer years of education (t=3.6, df = 64.1, p<.001)

### Clinical Disease Severity and Functional Status

Black individuals had higher CDR total (β = 1.21, SE= 0.57, p = .03) and higher CDR global (β = .65, SE= 0.24, p = .006) at their initial visit after adjusting for age, sex, education, and disease duration in linear regression models (Table 3). Black individuals also had a higher FAS total (β = 3.84, SE= 1.50, p = .01). We also observed several differences on FAS individual items (see Supplementary Table 1). Black individuals were more likely to have more difficulty with all but three items on the FAS. Items which did not differ by race include: 1) writing checks, paying bills, balancing checkbook; 2) shopping alone for clothes, household necessities, or groceries; and, 3) playing a game of skill, working on a hobby.

**Table 3:** Clinical Dementia Rating (CDR) Scale and Functional Assessment Scale (FAS) in FTD

| | bvFTD | | PPA | | bvFTD or PPA | | $\beta$ | Standard error | Odds Ratio (95% CI) | p |
| --- | --- | --- | --- | --- | --- | --- | --- | --- | --- | --- |
|  | White<br>(N = 1210) | Black<br>(N = 37) | White<br>(N = 1146) | Black<br>(N = 26) | White<br>(N = 2356) | Black<br>(N = 63) |  |  |  |  |
| CDRSUM<br>(mean (SD)) | 7.42<br>(4.49) | 8.57<br>(4.54) | 4.45<br>(4.27) | 5.31<br>(4.99) | 5.97<br>(4.63) | 7.22<br>(4.96) | 1.21 | 0.57 |  | .03 |
| CDRGLOB<br>(mean (SD)) | 1.29<br>(0.79) | 1.57<br>(0.84) | 0.83<br>(0.72) | 1.02<br>(0.91) | 1.07<br>(0.79) | 1.34<br>(0.91) | 0.65 | 0.24 | 1.20<br>(1.91, 3.03) | .006 |
| FAS<br>TOTAL<br>(mean (SD)) | 20.08<br>(8.22) | 24.38<br>(6.44) | 13.00<br>(9.98) | 15.76<br>(10.99) | 16.68<br>(9.76) | 20.80<br>(9.52) | 3.84 | 1.50 |  | 0.01 |

\*Statistical models were adjusted for age, sex, education, and disease duration.
Abbreviations: CDRSUM, CDR sum of boxes; CDRGLOB, CDR global.

### Neuropsychiatric Symptoms in FTD

As shown in Table 4, when compared to White individuals, Black individuals exhibited a higher prevalence of delusions, agitation, and depression on the NPI-Q (95% CI for odds ratio: delusions; 1.15-3.95, p = .01; agitation, 1.02-2.92; p = .04; depression: 1.06-2.94; p = 0.03), even after adjusting for age, sex, education, and CDR global. Similarly, in addition to prevalence of symptoms, we observed that Black individuals had greater symptom severity for delusions, agitation, and depression on each NPI-Q behavioral domain severity score (95% CI for odds ratio: delusions: 1.09-3.65; p =.02; agitation: 1.12-2.92; p = .01, depression: 1.00-2.54; p = .04). In contrast, Black individuals with FTD were less likely to exhibit apathy (95% CI for odds ratio: apathy: 0.32-0.94; p =.03) and showed a lower apathy severity (95% CI for odds ratio: apathy: 0.37-1.00; p = .048) compared to White individuals with FTD. There were no significant differences in NPI-Q total symptoms nor NPI-Q total severity between Black and White individuals. When stratifying the sample by bvFTD or PPA diagnosis we observe similar proportions and same direction of neuropsychiatric features across Black and White individuals (see Supplementary Table 2).

**Table 4:**
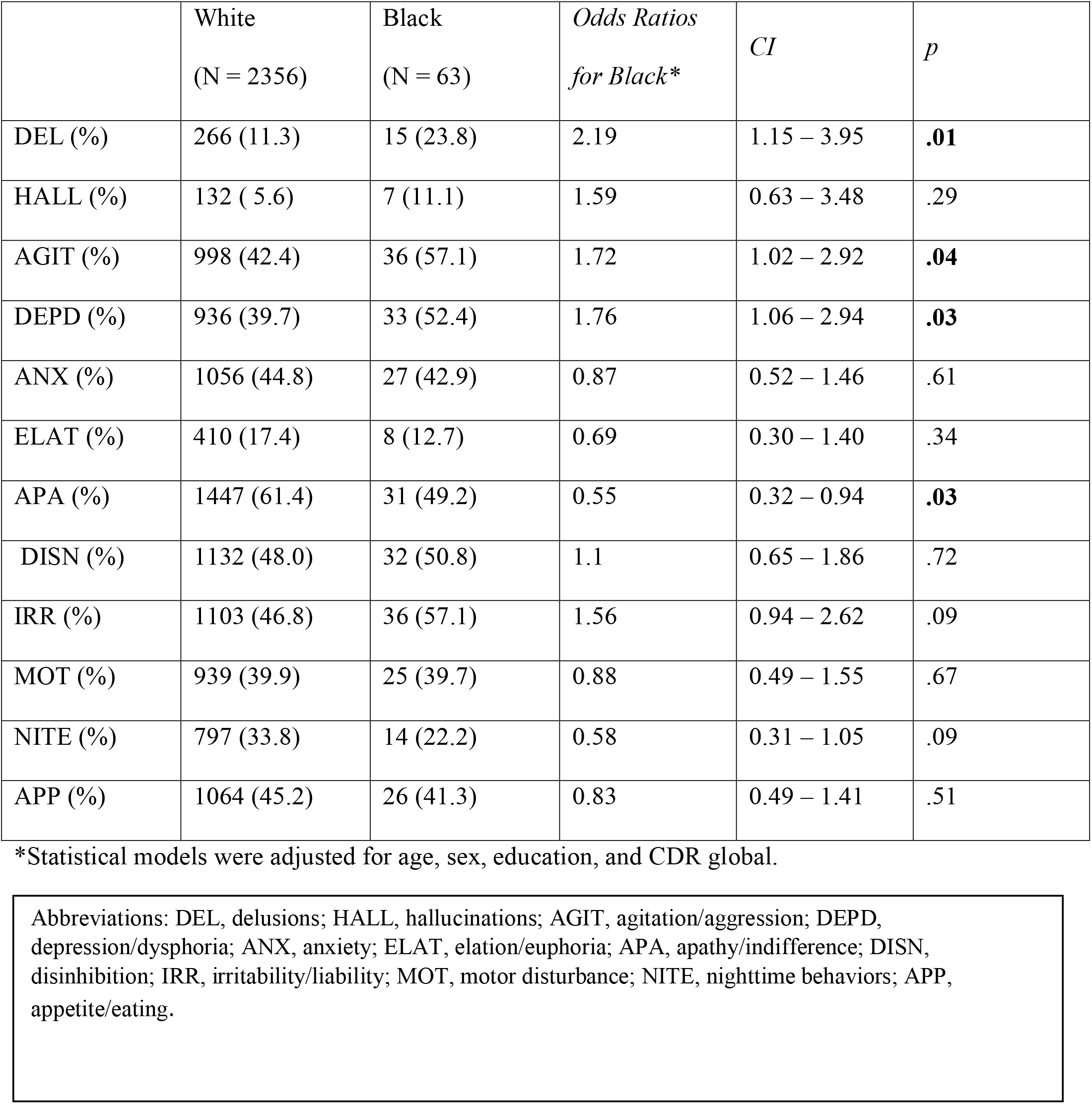
Prescence of neuropsychiatric symptoms in FTD

|  | White<br>(N = 2356) | Black<br>(N = 63) | *Odds Ratios*<br>*for Black\** | *CI* | *p* |
| --- | --- | --- | --- | --- | --- |
| DEL (%) | 266 (11.3) | 15 (23.8) | 2.19 | 1.15 – 3.95 | **.01** |
| HALL (%) | 132 ( 5.6) | 7 (11.1) | 1.59 | 0.63 – 3.48 | .29 |
| AGIT (%) | 998 (42.4) | 36 (57.1) | 1.72 | 1.02 – 2.92 | **.04** |
| DEPD (%) | 936 (39.7) | 33 (52.4) | 1.76 | 1.06 – 2.94 | **.03** |
| ANX (%) | 1056 (44.8) | 27 (42.9) | 0.87 | 0.52 – 1.46 | .61 |
| ELAT (%) | 410 (17.4) | 8 (12.7) | 0.69 | 0.30 – 1.40 | .34 |
| APA (%) | 1447 (61.4) | 31 (49.2) | 0.55 | 0.32 – 0.94 | **.03** |
| DISN (%) | 1132 (48.0) | 32 (50.8) | 1.1 | 0.65 – 1.86 | .72 |
| IRR (%) | 1103 (46.8) | 36 (57.1) | 1.56 | 0.94 – 2.62 | .09 |
| MOT (%) | 939 (39.9) | 25 (39.7) | 0.88 | 0.49 – 1.55 | .67 |
| NITE (%) | 797 (33.8) | 14 (22.2) | 0.58 | 0.31 – 1.05 | .09 |
| APP (%) | 1064 (45.2) | 26 (41.3) | 0.83 | 0.49 – 1.41 | .51 |

\*Statistical models were adjusted for age, sex, education, and CDR global.
Abbreviations: DEL, delusions; HALL, hallucinations; AGIT, agitation/aggression; DEPD, depression/dysphoria; ANX, anxiety; ELAT, elation/euphoria; APA, apathy/indifference; DISN, disinhibition; IRR, irritability/liability; MOT, motor disturbance; NITE, nighttime behaviors; APP, appetite/eating.

### Neuropsychiatric Symptoms in Normal Controls

The mean age in the sample of Black and White individuals in the normal control group (n=11,021) was 68.8 (SD= 10.8) years old. Mean years of education in the normal control group was 15.9 (SD=2.8) years. As shown in Table 2, there were more females in Black normal control group (*x*^2^=119.5, df = 1; p<.001) compared to White controls. Black normal control participants were also older (t = -2.2, df = 3489.6; p=.02) and had less education (t = 17.2, df = 2610.9; p<.001) than White normal control participants. Within the normal control group, Black individuals reported less depression, anxiety, agitation, and nighttime behaviors (95% CI for odds ratio: depression: 0.60-0.85; p < .001; anxiety: 0.61 – 0.92; p = .006; agitation: 0.56 – 0.99; p =.048; nighttime: 0.57 – 0.84; p <.001; see Table 5) compared to White individuals.

**Table 5:** Prescence of neuropsychiatric symptoms in healthy control participants

|  | White<br>(N = 9122) | Black<br>(N = 1899) | *Odds Ratios*<br>*for Black\** | *CI* | *p* |
| --- | --- | --- | --- | --- | --- |
| DEL (%) | 42 ( 0.5) | 9 ( 0.5) | 1.04 | 0.47 – 2.07 | .92 |
| HALL (%) | 14 ( 0.2) | 2 ( 0.1) | 0.64 | 0.10 – 2.35 | .56 |
| AGIT (%) | 360 ( 3.9) | 58 ( 3.1) | 0.75 | 0.56 – 0.99 | **.048** |
| DEPD (%) | 1033 (11.3) | 164 ( 8.6) | 0.72 | 0.60 – 0.85 | **<.001** |
| ANX (%) | 732 ( 8.0) | 116 ( 6.1) | 0.75 | 0.61 – 0.92 | **.006** |
| ELAT (%) | 47 ( 0.5) | 14 ( 0.7) | 1.38 | 0.72 – 2.48 | .31 |
| APA (%) | 246 ( 2.7) | 53 ( 2.8) | 1.01 | 0.73 – 1.36 | .96 |
| DISN (%) | 150 ( 1.6) | 26 ( 1.4) | 0.85 | 0.54 – 1.28 | .45 |
| IRR (%) | 796 ( 8.7) | 140 ( 7.4) | 0.83 | 0.69 – 1.00 | .06 |
| MOT (%) | 82 ( 0.9) | 11 ( 0.6) | 0.67 | 0.33 – 1.21 | .21 |
| NITE (%) | 867 ( 9.5) | 132 ( 7.0) | 0.69 | 0.57 – 0.84 | **<.001** |
| APP (%) | 392 ( 4.3) | 95 ( 5.0) | 1.09 | 0.85 – 1.37 | .49 |

\*Statistical models were adjusted for age, sex, education.

## DISCUSSION

We examined disparities in disease severity, functional impairment and neuropsychiatric symptoms comparing Black and White individuals from the NACC cohort clinically diagnosed with FTD. We observed that Black individuals with FTD had a higher prevalence and more severe delusions, agitation and depression compared to White individuals with FTD who demonstrated more apathy than Black individuals. White members of the normal control group were more likely to exhibit agitation, anxiety, depression and nighttime behavior compared to Black members of the normal control group. We also observed that Black individuals with FTD had higher levels of functional impairment at their initial visit compared to White individuals with FTD, even though they did not differ in disease duration. These findings suggest that there are racialized differences in neuropsychiatric symptom profiles and the extent of functional impairment at initial presentation for FTD.

Neuropsychiatric symptoms are exceedingly common in FTD.^3^ To our knowledge, this study is the first to demonstrate differences in neuropsychiatric profiles by race in FTD. A few studies have reported on racial differences in neuropsychiatric symptoms in other dementias such as AD^5,6^ and heterogenous sample of dementia.^17^ These studies are largely consistent with the findings in this present study whereby Black individuals are more likely to exhibit psychotic symptoms cluster,^5,8,9^ including delusions^5,17^ and agitation.^17^ Considering the diagnosis of FTD is based on clinical symptoms, racial disparities in the recorded presentation of Black individuals is concerning and may suggest that Black FTD patients may be at particular risk for misdiagnosis if their symptom profile does not fit within the current clinical criteria.^3^ We also observed higher rates of depression, as well as lower rates of apathy, on the NPI-Q in Black individuals with FTD relative to White individuals with FTD, which is inconsistent with existing literature showing lower prevalence of depressed mood and dysphoria in Black older adults with and without dementia^18,19^ and studies showing no racial differences in affective symptoms.^17,20^ The lower prevalence of apathy in our Black sample may be related to sex and gender effects on reporting of affective symptoms, as older men have been shown to present more commonly with apathy without depressed mood,^21,22^ and our White FTD sample included significantly more men. Apathy was also most commonly reported in the White bvFTD subgroup, which had the highest proportion of male participants of all study subgroups. Due to the small size of Black individuals with FTD, our findings are not valid estimates of symptom prevalence for Black indivduals with FTD. Further, differences in reported rates of depressed mood and apathy may reflect a high degree of phenomenological overlap between apathy and depression, which share multiple features and are commonly misdiagnosed even by expert clinicians.^23^ Future work should consider utilizing more specific measures of depression and apathy to disentangle potential racial differences in affective symptom presentation in dementia.

We also observed that Black individuals with FTD were more likely to endorse depression on the NPI-Q. The literature on differences in rate of depression in Black and White individuals diagnosed with dementia is limited to a small number of studies that report mixed results such as no racial differences in affective symptoms^17,20^ or greater frequency of depression in White individuals compared to Black individuals with dementia.^8^ One possibility for our finding is the way in which depression was ascertained on the NPI-Q (e.g. “Does the patient act as if he or she is sad or in low spirits? Does he or she cry?”). Depression is often difficult to distinguish from apathy,^24^ a core feature of bvFTD^3^ and it is possible that Black care partners of persons with FTD are less likely to reocognize apathy and rather endorse a more familiar symptom such as depression. Future work should consider utilizing more specific or relevant measures of depression symptoms to confirm our findings and better understand that which underlies potential differences in symptoms of depression by race.

We also observed that Black individuals demonstrated greater levels of functional impairment compared to White counterparts even though disease duration did not differ between racial groups. Clinician ratings on the CDR may be influenced by implicit bias, but Black individuals were also more impaired on the FAS which is a care partner rated questionnaire suggesting the severity of functional impairment in Black individuals is not entirely biased by clinician rating. This is in line with prior research showing that Black individuals with dementia show worse performance on measures of cognitive and everyday functioning compared to White individuals with dementia.^11,25^ One possibility is that Black individuals may delay seeking medical attention for more subtle symptoms associated with cognitive and behvavioral decline.^17,26^ Studies suggest that social attitudes in the Black community about aging may cause a delay in seeking medical treatment;^27^ yet, it may be that certain neuropsychiatric symptoms such as delusions and agitation which can be alarming and pose a danger to patients and caregivers are difficult to overlook leading to Black caregivers to seek medical attention.

Importantly, Black individuals were considerably underrepresented in this dataset, comprising only 2.5% of our study sample. Out of 36 ADRCs contributing data to NACC, 14 (38.9%) did not include any Black individuals with FTD. Underrepresentation of Black and patients from other minoritized groups impedes the accuracy of population level research and limits our ability to identify the extent and etiologies of disparities in FTD. The causes of racial disparities in FTD prevalence and symptoms are likely complex and may be interrelated to the causes of racial disparities in healthcare utilization. There are likely multiple systemic and structural barriers that limit equitable access to health information (such as on the signs and symptoms of atypical dementias) and the specialized healthcare^28,29^ needed for timely diagnosis and management of FTD. Even when health information and healthcare resources are accessible, lack of trust in health systems^30^ that has been perpetuated by a history of institutional racism^31^ and higher costs associated with the dementia care received by Black individuals may deter healthcare seeking behavior.^32^ In this context, Black individuals with FTD may choose to delay medical care for the neuropsychiatric symptoms they experience, relying instead on faith-based coping strategies and support from loved ones.^10^ To date, the enumeration of racial disparities in ADRD care utilization, in general, demonstrate mixed results.^33–35^ Research on the root causes of racial disparities in context-level determinants of healthcare access and healthcare utilization is critically needed. This is especially so because FTD specialist care is often linked to research opportunities as well as diagnostic recognition in registries such as the NACC, which in turn informs population level understanding of the prevelance and symptomatology of FTD.

This study should be interpreted in light of several limitations. Dementia-related behaviors are often measured using caregiver report and differences in caregiver characteristics such as sex, race and education may result in differential reporting of behaviors.^36,37^ The CDR is a clinician interpretation of the individual’s level of functioning and therefore may be subject to unconscious bias exacerbated in the context of inadequate normative data in Black samples.^38^ Limitations in power did not allow us to examine differences on neuropsychological measures of cognition but as more data is collected, this may be an important area for future study. We also relied on clinical diagnosis in NACC and confirming these findings in a pathologically defined sample would be an important next step. We analyzed cross-sectional data from initial visit as longitudinal data were limited in Black individuals and therefore, we cannot attest to possible longitudinal changes in neuropsychiatric profile and functional decline. We also recognize that our analysis, situated in investigating differences through a Black-White racial binary is just one of many important comparisons; understanding disparities and the reasons that underlie underrepresentation of minoritized patients in clinical data and participants in clinical research is a much broader gap in current knowledge. It remains to be determined if the disparities observed in AD and other forms of dementia are the same as those in FTD. As FTD is rarer than AD, any disparities caused by decreased awareness of dementias or caused by decreased access to care might be more pronounced in FTD compared to AD. New knowledge about facilitators and barriers can help researchers develop more effective strategies for engaging currently underrepresented individuals.

With these caveats in mind, this study suggests that there are differences in profiles of neuropsychiatric symptoms, greater disease severity and lower scores on functional measures in Black and White individuals diagnosed with FTD. These findings corroborate existing research that suggest there are disparities in dementia symptoms and functional impairment in Black and White individuals living with ADRD. The reasons for these differences and their implications remain poorly understood. Future work must address disparities in FTD and the systemic and structural determinants that drive them. Efforts to promote equitable access to health care and enrollment in clinical research should be prioritized.

## Data Availability

Qualified researchers may obtain access to all de-identified data in the NACC registry used for this study (http://www.naccdata.org).

## ACKNOWLEDGEMENTS

The NACC database is funded by NIA/NIH Grant U24 AG072122. NACC data are contributed by the NIA-funded ADRCs: P30 AG062429 (PI James Brewer, MD, PhD), P30 AG066468 (PI Oscar Lopez, MD), P30 AG062421 (PI Bradley Hyman, MD, PhD), P30 AG066509 (PI Thomas Grabowski, MD), P30 AG066514 (PI Mary Sano, PhD), P30 AG066530 (PI Helena Chui, MD), P30 AG066507 (PI Marilyn Albert, PhD), P30 AG066444 (PI John Morris, MD), P30 AG066518 (PI Jeffrey Kaye, MD), P30 AG066512 (PI Thomas Wisniewski, MD), P30 AG066462 (PI Scott Small, MD), P30 AG072979 (PI David Wolk, MD), P30 AG072972 (PI Charles DeCarli, MD), P30 AG072976 (PI Andrew Saykin, PsyD), P30 AG072975 (PI David Bennett, MD), P30 AG072978 (PI Neil Kowall, MD), P30 AG072977 (PI Robert Vassar, PhD), P30 AG066519 (PI Frank LaFerla, PhD), P30 AG062677 (PI Ronald Petersen, MD, PhD), P30 AG079280 (PI Eric Reiman, MD), P30 AG062422 (PI Gil Rabinovici, MD), P30 AG066511 (PI Allan Levey, MD, PhD), P30 AG072946 (PI Linda Van Eldik, PhD), P30 AG062715 (PI Sanjay Asthana, MD, FRCP), P30 AG072973 (PI Russell Swerdlow, MD), P30 AG066506 (PI Todd Golde, MD, PhD), P30 AG066508 (PI Stephen Strittmatter, MD, PhD), P30 AG066515 (PI Victor Henderson, MD, MS), P30 AG072947 (PI Suzanne Craft, PhD), P30 AG072931 (PI Henry Paulson, MD, PhD), P30 AG066546 (PI Sudha Seshadri, MD), P20 AG068024 (PI Erik Roberson, MD, PhD), P20 AG068053 (PI Justin Miller, PhD), P20 AG068077 (PI Gary Rosenberg, MD), P20 AG068082 (PI Angela Jefferson, PhD), P30 AG072958 (PI Heather Whitson, MD), P30 AG072959 (PI James Leverenz, MD).

## Footnotes

i Some participants received an FTD diagnosis only after their first NACC visit, meaning that their “initial visit” as defined in this current study was not their first NACC visit. The patient’s first NACC visit was not their initial visit for 17.4% of Black participants and 13.2 % of White participants in our final sample.

