## Supplemental File 1 for "Differences in Neuropsychiatric Features in Black and White Individuals Diagnosed with Frontotemporal Degeneration"

Supplementary Table 1: Functional Assessment Scale (FAS) Items in FTD

| | White (N = 1669) | Black (N = 41) | *Odds Ratios for Black* | *CI* | *p* |
|---|---|---|---|---|---|
| BILLS (%) | 893 (53.5) | 27 (65.9) | 1.6 | 0.83 – 3.19 | .17 |
| TAXES (%) | 971 (58.2) | 33 (80.5) | 2.8 | 1.33 – 6.61 | **.01** |
| SHOPPING (%) | 589 (35.3) | 21 (51.2) | 1.81 | 0.95 – 3.45 | .07 |
| GAMES (%) | 473 (28.3) | 15 (36.6) | 1.39 | 0.71 – 2.66 | .32 |
| STOVE (%) | 432 (25.9) | 18 (43.9) | 2.19 | 1.13 – 4.16 | **.02** |
| MEALPREP (%) | 623 (37.3) | 23 (56.1) | 2.13 | 1.13 – 4.07 | **.02** |
| EVENTS (%) | 426 (25.5) | 22 (53.7) | 3.09 | 1.63 – 5.90 | **.001** |
| PAY ATTN (%) | 324 (19.4) | 15 (36.6) | 2.32 | 1.17 – 4.45 | **.01** |
| REM DATES (%) | 525 (31.5) | 22 (53.7) | 2.41 | 1.27 – 4.60 | **.01** |
| TRAVEL (%) | 801 (48.0) | 30 (73.2) | 2.68 | 1.35 – 5.70 | **.01** |

*Statistical models were adjusted for age, sex, education, and disease duration.
