## Supplemental File 2 for "Differences in Neuropsychiatric Features in Black and White Individuals Diagnosed with Frontotemporal Degeneration"

Supplementary Table 2: CDR and NPI by FTD phenotype

| | **bvFTD** | | **PPA** | |
|---|---|---|---|---|
| | White | Black | White | Black |
| n | 1210 | 37 | 1146 | 26 |
| CDRGLOB (mean (SD)) | 1.29 (0.79) | 1.57 (0.84) | 0.83 (0.72) | 1.02 (0.91) |
| DEL = 1 (%) | 189 (15.6) | 10 (27.0) | 77 ( 6.7) | 5 (19.2) |
| HALL = 1 (%) | 103 ( 8.5) | 5 (13.5) | 29 ( 2.5) | 2 ( 7.7) |
| AGIT = 1 (%) | 628 (51.9) | 23 (62.2) | 370 (32.3) | 13 (50.0) |
| DEPD = 1 (%) | 478 (39.5) | 18 (48.6) | 458 (40.0) | 15 (57.7) |
| ANX = 1 (%) | 576 (47.6) | 15 (40.5) | 480 (41.9) | 12 (46.2) |
| ELAT = 1 (%) | 280 (23.1) | 6 (16.2) | 130 (11.3) | 2 ( 7.7) |
| APA = 1 (%) | 924 (76.4) | 23 (62.2) | 523 (45.6) | 8 (30.8) |
| DISN = 1 (%) | 781 (64.5) | 22 (59.5) | 351 (30.6) | 10 (38.5) |
| IRR = 1 (%) | 647 (53.5) | 24 (64.9) | 456 (39.8) | 12 (46.2) |
| MOT = 1 (%) | 644 (53.2) | 18 (48.6) | 295 (25.7) | 7 (26.9) |

| | | | | |
|---|---|---|---|---|
| NITE = 1 (%) | 508 (42.0) | 12 (32.4) | 289 (25.2) | 2 ( 7.7) |
| APP = 1 (%) | 680 (56.2) | 18 (48.6) | 384 (33.5) | 8 (30.8) |
